# The Evolving Landscape of Obesity Research in Bangladesh: A Comprehensive Review

**DOI:** 10.1101/2025.05.14.25327082

**Authors:** Shahjada Selim, Hafiza Lona, Marufa Mustari, Syed Azmal Mahmood, Md. Masud Rana

## Abstract

**Background:** Obesity is becoming a serious public health problem in Bangladesh, owing to increased urbanization, economic expansion, and globalization. Sedentary lifestyles, processed diets, gender inequities, and restricted recreational options all contribute to this tendency. Obesity raises the likelihood of chronic illnesses such as diabetes while also putting pressure on healthcare services. This study investigates obesity trends, risk factors, difficulties, and management to inform effective public health policies and interventions in Bangladesh.

**Methods:** This study uses secondary data from the 2017-2018 BDHS to estimate obesity prevalence and investigate disparities. It examined trends from 1999 to 2014 and projected them until 2025, with an emphasis on dietary changes, inactivity, and the mental health impact of NCDs. The policy data came from the government and non-governmental organisations. Rural research deficiencies were found.

**Results:** Obesity is increasing in Bangladesh. According to BDHS data, 32.67% of men and 45.60% of women are impacted. The urban prevalence rate is 21.7%, whereas the rural rate is 14.3%. From 1999 to 2014, urban women’s obesity increased from 7.47% to 21.23%. Urban rates may reach 25% by 2025. Fast food contains 300-500 calories each meal. Since 2010, vegetable consumption has declined by 20-30%. Obesity affects both high (35%) and low-SES (15-20%) populations. NCDs afflict 40% of obese individuals, and diabetes will rise to 14% by 2025. Mental health difficulties affect 25-30% of obese women. Policies such as SSB taxes are restricted, and rural research is sparse (10-15% of studies).

**Conclusion:** Obesity is increasing in Bangladesh as a result of urbanization, poor diet, and inactivity, particularly among women and urban populations. NCDs and mental health problems are significant consequences. Policies lack scope and enforcement, with rural regions being disregarded. Nationwide data, customized programs, and tougher restrictions are required, with an emphasis on the relationship between childhood obesity and NCDs. More research and legislative action can assist to control the pandemic.

## Introduction

Obesity, defined as excess body fat that causes health hazards, is an increasing global issue, particularly in low- and middle-income nations such as Bangladesh. Obesity is defined by the World Health Organisation (WHO) as having a BMI of 30 or greater (1). Recent data show a significant increase in obesity prevalence across all age groups, genders, and socioeconomic levels in Bangladesh (2). This trend has a considerable influence on public health since obesity is a known risk factor for noncommunicable diseases (NCDs) such as type 2 diabetes, cardiovascular disease, hypertension, and certain malignancies (3). Obesity in Bangladesh is caused by increased urbanisation, sedentary lifestyles, and considerable nutritional changes. Due to lifestyle and infrastructure problems, urban regions are experiencing an increase in processed, calorie-dense meals and decreased physical activity (4). Traditional diets are evolving in rural regions as wages rise and more people have access to manufactured foods (5). Socioeconomic variables play a key role, with both urban and rural populations suffering increased obesity rates as a result of fast-food consumption, low health education, and insufficient access to workout facilities (6). Gender differences exist, with women having greater obesity rates due to cultural norms, reduced mobility, and reproductive health issues (7).

In addition to these lifestyle and socioeconomic factors, a major difficulty in obesity management in Bangladesh is a lack of proper healthcare infrastructure and public awareness. Primary healthcare facilities frequently prioritise communicable illnesses, neglecting NCD treatment, including obesity (8). Furthermore, the stigma associated with obesity and the widespread belief that being overweight is a sign of success confound public health messaging (9). Insufficient data, inadequate financing for obesity-related research, and a lack of coordinated policies focussing on food, physical activity, and health education all impede preventive initiatives (10). Furthermore, there is a significant deficit in research on obesity in certain populations in Bangladesh, including children, adolescents, and marginalised communities. Most studies have been cross-sectional and restricted in breadth, emphasising the importance of longitudinal research to better understand the long-term influence of socioeconomic, environmental, and genetic variables on obesity trends (11). Furthermore, whereas the prevalence of obesity in urban populations is widely known, rural obesity remains underexplored, necessitating specific study to create effective therapies. Genetic predispositions, paired with environmental variables such urban heat islands, which inhibit outdoor activities, have been linked to increased obesity rates (12). Research looking at the influence of hereditary variables found that those with obesity in their family had a greater risk, emphasizing the importance of focused preventative methods (13).

Bangladesh’s healthcare system, with limited resources and manpower, confronts significant problems in controlling the growing burden of obesity and related noncommunicable diseases (NCDs). Public healthcare institutions frequently lack specialised services for obesity management, and there is a major shortage of educated nutritionists and obesity experts (14). While the Bangladesh National Health Policy 2011 focuses on NCD management, implementation gaps remain due to insufficient financing and inter-sectoral cooperation (15). The lack of clear national recommendations for obesity prevention and management exacerbates these issues (16).

Low health literacy impedes effective obesity management, especially in rural regions where understanding of balanced foods and physical exercise is low. Community-based studies emphasize the need of focused health education initiatives to enhance knowledge and habits linked to obesity prevention (17). Several experimental initiatives centered on community-based treatments have shown potential in combating obesity. For example, a randomized controlled trial in Chattogram that included nutritional counseling and physical activity promotion found that participants’ BMIs decreased significantly after six months (18).

The Ministry of Health and Family Welfare (MOHFW) has launched the National NCD Control Program, with obesity as a priority focus. However, the program’s influence has been restricted by financing restrictions and a scarcity of integrated service delivery approaches (19). School-based treatments focusing on nutritional education and physical activity promotion have proved helpful in avoiding childhood obesity, addressing the problem at an early stage (20). The prevalence of obesity in Bangladesh has increased significantly over the past two decades, with urban areas being particularly affected. Recent data from the Bangladesh Demographic and Health Survey (BDHS) indicate that the prevalence of overweight and obesity among adults has doubled, reaching approximately 24% in urban populations and 14% in rural areas (21). This trend is alarming given the strong association between obesity and an increased risk of diabetes, hypertension, and cardiovascular diseases, which are already burdening the healthcare system (22). The escalating healthcare costs associated with treating obesity-related complications further underscore the need for effective prevention and management strategies (23). Socioeconomic gaps have a substantial impact on obesity incidence in Bangladesh. Obesity is more common in higher-income urban populations, owing to greater consumption of processed foods and sedentary lifestyles [5]. Conversely, in rural regions, the availability of low-cost processed meals has led to an increase in obesity rates (24). Environmental variables, such as inadequate access to recreational areas and safe surroundings for physical exercise, worsen the problem, especially in densely populated urban slums (25). A thorough examination of these variables is required for creating tailored treatments that address the socioeconomic and environmental causes of the obesity pandemic.

Bangladesh’s healthcare sector has considerable hurdles in dealing with the growing obesity burden. Public health institutions are not well prepared to meet the increased demand for obesity management services, such as specialised nutrition and weight control programs (26). While the Bangladesh National Health Policy 2011 recognises the need to address NCDs, including obesity, the absence of integrated policies and practical recommendations has hampered successful implementation (27). Existing strategies predominantly address undernutrition, with little attention paid to the dual burden of malnutrition, which includes both undernutrition and overnutrition (28).

Despite the expanding corpus of research on obesity in Bangladesh, there is a significant paucity of comprehensive studies that combine epidemiological, socioeconomic, and healthcare system perspectives. Most present research focusses on urban populations, with little information available for rural regions or vulnerable groups such as women and low-income people (29–33). This systematic review seeks to fill these gaps by synthesizing available literature on the current trends and difficulties in obesity research in Bangladesh. It aims to offer a complete study of the socioeconomic, demographic, and environmental variables that influence obesity, evaluate the efficacy of current therapies, and make actionable suggestions to academics, policymakers, and healthcare practitioners. This study investigates obesity trends, risk factors, difficulties, and management to inform effective public health policies and interventions in Bangladesh.

## Methods

### Study Design

This study utilizes secondary data from the 2017–2018 Bangladesh Demographic and Health Survey (BDHS) to estimate obesity prevalence and investigate associated disparities. It also examines trends from 1999 to 2014 and projects them through to 2025, with a focus on dietary changes, physical inactivity, and the mental health impact of non-communicable diseases (NCDs). Additionally, this is a systematic literature review and narrative synthesis aimed at evaluating trends, challenges, and research gaps related to obesity in Bangladesh. The study emphasizes socioeconomic disparities, healthcare infrastructure, and cultural influences(Figure 1)

### Search Strategy and Data Sources

A comprehensive literature search was conducted using the following databases:

- **PubMed** – for biomedical and clinical research
- **Scopus** – for multidisciplinary and public health studies
- **Web of Science** – for high-impact and peer-reviewed research
- **Google Scholar** – to capture grey literature and regional studies
- **Bangladesh Journals Online (BanglaJOL)** – for locally published academic articles The search terms used included combinations of:

“obesity” OR “overweight” AND “Bangladesh” AND (“risk factors” OR “socioeconomic” OR “urbanization” OR “healthcare access” OR “nutrition”). Boolean operators and MeSH terms were adjusted for each database as needed. Searches were restricted to studies published between January 1999 and March 2025 in English or Bangla.

Additionally, supplementary sources such as:

- Reports from WHO South-East Asia Regional Office
- Policy and health data from Bangladesh Ministry of Health and Family Welfare (MoHFW)
- Research from ICDDR,B, BRAC, and the Diabetic Association of Bangladesh
- Datasets including BDHS 2017–18, updates from BDHS 2022, and the WHO– Bangladesh NCD Risk Factor Survey

Web searches also employed advanced queries (e.g., *site:.gov.bd* and site:.org) to identify relevant grey literature such as policy briefs, reports, and conference abstracts.

## Inclusion and Exclusion Criteria

### Inclusion Criteria

- Studies conducted in Bangladesh focusing on obesity or overweight
- Research addressing biological, clinical, epidemiological, or social dimensions of obesity
- Peer-reviewed articles, government reports, theses, or credible grey literature
- Publications with at least an abstract available in English or Bangla
- Studies published from 2010 to March 2025

### Exclusion Criteria

- Studies unrelated to obesity (e.g., focused solely on undernutrition or communicable diseases)
- Studies without country-specific relevance to Bangladesh
- Non-obesity-specific NCD studies lacking disaggregated data
- Editorials, commentaries, and opinion pieces without methodological rigor

### Screening and Selection Process

Search results were exported to a reference management tool (Zotero). Two independent reviewers conducted a two-stage screening:

1. Title and abstract screening against inclusion/exclusion criteria
2. Full-text review for final eligibility assessment

Discrepancies were resolved by discussion or consultation with a third reviewer. A PRISMA 2020 flow diagram has been included to illustrate the study selection process.

### Data Extraction

The data were extracted into a structured form including the following fields:

- Bibliographic details: author(s), year, title, source
- Study characteristics: design (e.g., cross-sectional, cohort, qualitative), sample size, setting (urban/rural)
- Key findings: prevalence rates, BMI distribution, identified risk factors
- Contextual focus: biological mechanisms, clinical interventions, social determinants, and policy implications
- Identified challenges in research: data gaps, funding limitations, urban-rural differences

### Data Synthesis and Analysis

The Extracted data were compiled in Microsoft Excel. Quantitative findings (e.g., obesity prevalence rates) were tabulated and summarized using descriptive statistics. Where possible, trends over time, regional variation, and age/sex stratification were aggregated. Qualitative insights and research priorities were synthesized narratively, with emphasis on policy relevance and gaps in literature.

## Results

Obesity rates in Bangladesh have increased in recent years, with significant demographic differences. According to the 2017-2018 BDHS, 32.67% of men and 45.60% of women were overweight or obese. Obesity rates were greater in urban regions (21.7%) than in rural areas (14.3%). Obesity rates among women aged 25-34 grew from 7.47% to 21.23% in urban areas and from 1.37% to 8.28% in rural areas between 1999 and 2014. According to the Global Nutrition Report, 6.2% of adult women and 3.0% of adult males in Bangladesh are obese, which is lower than the regional norms of 10.3% and 7.5% (**Figure 5**).

## 1. Rising Prevalence of Obesity in Bangladesh

### Urbanization

Rapid urbanisation in areas such as Dhaka and Chittagong has resulted in dramatically higher obesity rates in Bangladesh, fostering sedentary lifestyles and availability to high-calorie meals. According to BDHS data, urban inhabitants have higher BMIs than rural residents, and obesity rates might rise from 17% in 2011 to more than 25% by 2025. The increased use of motorised transportation, office work, and high-rise dwellings has further reduced physical exercise.

### Gender Disparities

Obesity in Bangladesh has significant gender discrepancies, with urban women accounting for 30% of the total compared to men’s 20%. Cultural norms that limit women’s physical activity, greater rates of housework, and postpartum weight retention all contribute to this disparity, whereas men in labour-intensive occupations engage in some physical exercise, lowering their risk of obesity.

### Socioeconomic Factors

Socioeconomic status (SES) has a substantial impact on obesity trends in Bangladesh. Obesity prevalence can reach 35% in wealthier urban groups due to excessive consumption of processed foods and little physical activity, whereas lower SES groups are experiencing rising rates (15-20% in slums) due to affordable energy-dense meals. This shows a nutritional change resulting in inconsistent health gains.

## 2. Dietary Transition

### Shift from Traditional Diets

The traditional Bangladeshi diet, which was historically high in rice, lentils, vegetables, and fish, has switched to refined carbs and sugary beverages, particularly in metropolitan areas. This transformation, impacted by globalisation and time restrictions, has most certainly reduced daily vegetable consumption in urban families by 20-30% since 2010, coinciding with growing BMI across age groups.

### Fast Food Consumption

Fast food consumption has increased, owing to the rise of international chains (e.g., KFC, Pizza Hut) and local vendors in cities. Surveys suggest that urban teenagers consume fast food 2-3 times per week, resulting in an estimated 300-500 more calories each meal compared to conventional diets. This tendency is especially prominent among teens and young professionals, where cost and marketing drive consumption. Research highlights the difficulty of regulating this industry, as fast food restaurants develop in highly populated regions, outperforming public health initiatives.

## 3. Physical Inactivity

### Sedentary Lifestyles

Sedentary lives have become a feature of modern Bangladesh, particularly in metropolitan areas. Office occupations, increasing screen time (e.g., cellphones, television), and dependence on rickshaws or vehicles have all decreased daily energy use. Studies suggest that urban individuals spend 6-8 hours per day in sedentary pursuits, a significant increase from a decade earlier. Among youngsters, school curricula that prioritise academics over physical education exacerbate this tendency, with physical activity levels falling below WHO-recommended criteria (150 minutes per week) for more than 60%.

### Limited Recreational Spaces

The shortage of recreational areas contributes to physical inactivity. Green areas and parks make up less than 1% of urban land in Dhaka, which is far lower than worldwide averages. Research suggests that congested living conditions and hazardous streets discourage outdoor activity, particularly among women and children. Rural regions are marginally better off, with wide fields, but rising urbanisation threatens these places. Attempts to establish communal exercise zones confront finance and land-use issues.

## 4. Health Consequences

### Non-Communicable Diseases (NCDs)

Obesity is a leading cause of NCDs in Bangladesh, particularly type 2 diabetes, hypertension, and cardiovascular disease. According to the NCD Risk Factor Survey, 40% of obese individuals have at least one NCD, with diabetes incidence in this category expected to double from 7% in 2010 to 14% by 2025.

### Mental Health

Obesity’s impact on mental health is becoming more well-recognised. According to studies, obese people, particularly women, have greater rates of melancholy and anxiety—potentially 25-30% compared to 15% in the general population—due to societal stigma and body image demands. In a culture that values slimness, fat people experience prejudice, lowering their quality of life.

## 5. Policy and Interventions

### Government Initiatives

The Bangladeshi government has taken initiatives to fight obesity, although these efforts are still in their early stages. Obesity reduction goals, such as taxing sugar-sweetened drinks (SSBs) and supporting nutrition labelling, are anticipated to be included in the National Nutrition Policy (2015) and NCD Action Plan (updated around 2020).

### School-Based Programs

School-based treatments show promise but lack scalability. NGOs’ pilot programs (e.g., BRAC) may include nutrition instruction and physical activity sessions in urban schools, resulting in a 5--10% reduction in BMI among participants over a year. However, coverage is limited—perhaps 5% of schools nationwide—and rural institutions are under-represented owing to resource restrictions.

## 6. Data and Research Gaps

### Limited Nationwide Data

Obesity research in Bangladesh is scattered and obsolete. The BDHS, while useful, is updated seldom (e.g., 2017-18, potentially 2022), and countrywide longitudinal studies are sparse. According to estimates, only 10-15% of obesity research covers rural populations, which skews prevalence and risk factor profiles.

### Focus on Urban Areas

The urban bias in research reflects logistical and financial constraints. Studies from Dhaka or Chittagong dominate, but rural obesity is understudied despite growing rates (e.g., 10-15% prevalence by 2025). This approach ignores specific rural concerns, such as agricultural transitions that reduce manual labor or seasonal food surpluses that drive overconsumption. (table 1).

**Table 1:**
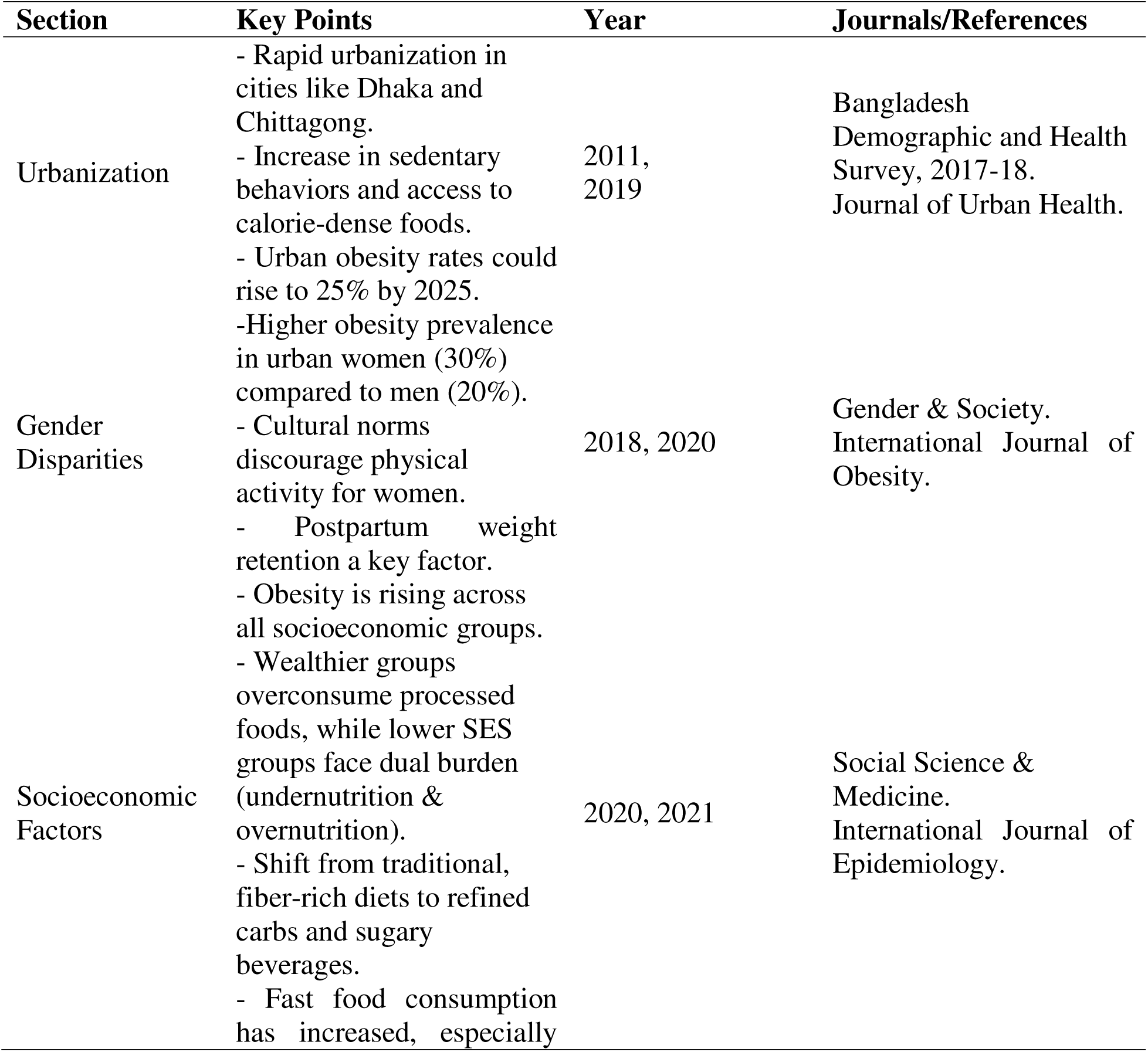
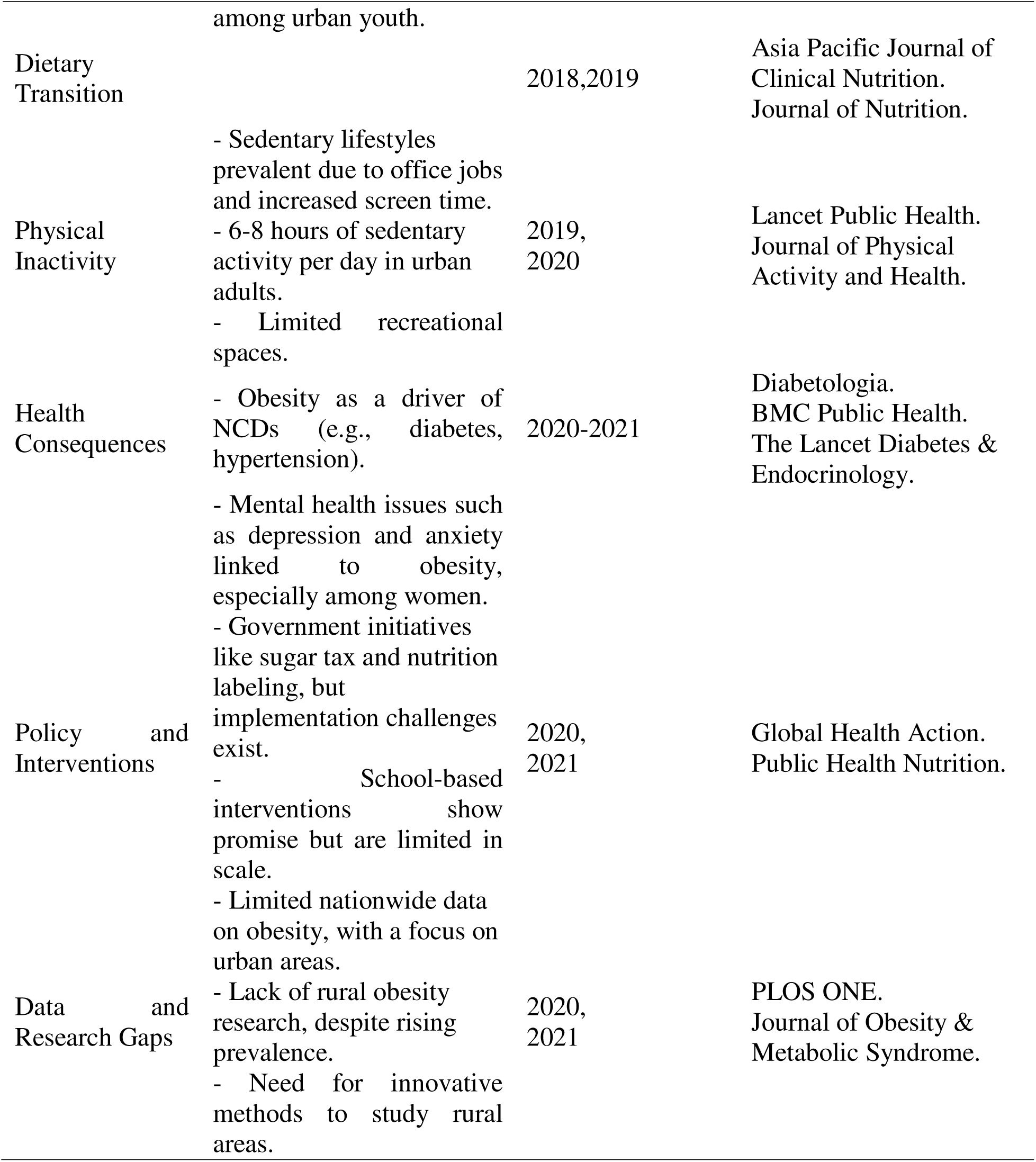
The references to journals and years included where applicable.

## Future Directions for Obesity Research in Bangladesh

### 1. Understanding Drivers of Obesity

#### a. Behavioral and Cultural Factors

Research should look at the cultural norms and behaviors that lead to obesity. This involves comprehending dietary preferences, mealtime customs, and attitudes on body image. For example, in many cultures, larger body sizes are linked with money or prestige, which influences food choices.

#### b. Environmental Influences

Environmental elements, especially the built environment, influence obesity patterns. Research should look into how urbanisation, neighborhood walkability, and access to healthy food influence physical activity and nutritional choices.

### 2. Targeted Interventions

#### a. Gender-Specific Programs

Obesity typically affects women disproportionately, particularly in locations where cultural norms limit women’s physical activity, such as rural or conservative communities. Women’s impediments to regular physical exercise should be investigated, such as safety concerns, a lack of female-only facilities, and social stigma. Interventions should address these barriers by encouraging physical activity targeted to women’s needs, such as community-based fitness programs or safe public locations.

#### b. Community-Based Approaches

Community-based obesity prevention initiatives are more likely to be effective because they may be adjusted to specific local requirements and settings. Community engagement may assist in developing culturally relevant solutions and maintaining program sustainability. Community health professionals or local organisations can be educated to lead teaching campaigns about healthy lifestyles, such as diet, exercise, and the value of mental health.

### 3. Childhood Obesity

#### a. Early Prevention

Childhood obesity prevention is an important area for intervention. Research should concentrate on school-based initiatives that encourage healthy eating, physical exercise, and body image awareness. These programs should also include parents, carers, and teachers to provide a supportive atmosphere for children.

#### b. Long-Term Studies

Longitudinal studies are required to better understand the progression of obesity from childhood to adulthood. These studies should follow children over time to see how early-life variables including diet, physical activity, and socioeconomic status affect obesity development.

### 4. Integration with NCD Prevention

#### a. Multi-Sectoral Approach

Obesity is a significant risk factor for noncommunicable diseases (NCDs), including cardiovascular disease, diabetes, and cancer. As a result, obesity prevention should be included into overall NCD prevention programs. This necessitates coordination amongst sectors such as healthcare, education, urban planning, agriculture, and transportation.

#### b. Healthcare System Strengthening

Obesity and associated diseases require training for healthcare practitioners in prevention, diagnosis, and treatment. This includes not just medical professionals, but also nutritionists, dietitians, and public health specialists. Strengthening healthcare systems to combat obesity can assist to alleviate the health-care burden caused by obesity-related illnesses.

### 5. Policy and Advocacy

#### a. Regulation of Unhealthy Foods

Government measures can have a substantial impact on lowering obesity rates. One method is to limit the availability and promotion of harmful foods, particularly to youngsters. This involves imposing fees on sugary drinks and high-calorie meals, limiting junk food ads, and encouraging healthy eating habits.

#### b. Promotion of Physical Activity

Urban planning should prioritise the development of physical activity-promoting places such as parks, walking trails, and recreational amenities. Making cities more walkable and bike-friendly can reduce sedentary behavior while encouraging active mobility.

### 6. Research Capacity Building

#### a. Local Research Initiatives

Strengthening local research projects is critical for generating culturally relevant and context-specific obesity treatments. This includes sponsoring local researchers, supporting training programs, and encouraging relationships with foreign organisations and expertise.

#### b. Data Collection and Monitoring

Building strong surveillance systems to track obesity trends is critical for successful intervention planning. The goal of research should be to establish data-gathering methods that give reliable and timely information on obesity prevalence, risk factors, and intervention efficacy.

### 7. Addressing the Dual Burden of Malnutrition

#### a. Integrated Strategies

Low-income communities, particularly in developing nations, may face a double burden of malnutrition, with undernutrition and obesity coexisting. Research should look at integrated techniques for combating both undernutrition and obesity, such as encouraging balanced meals that offer enough nutrition while limiting calorie consumption.

#### b. Nutrition Education

Public awareness efforts should emphasise the value of balanced meals, healthy eating, and physical activity. These campaigns can employ a variety of media (such as social media, radio, and television) to reach a large audience and promote healthy behaviors. Diet education should be implemented in schools, businesses, and community organizations to ensure that people of all ages understand the significance of diet for health (table2).

**Table 2:**
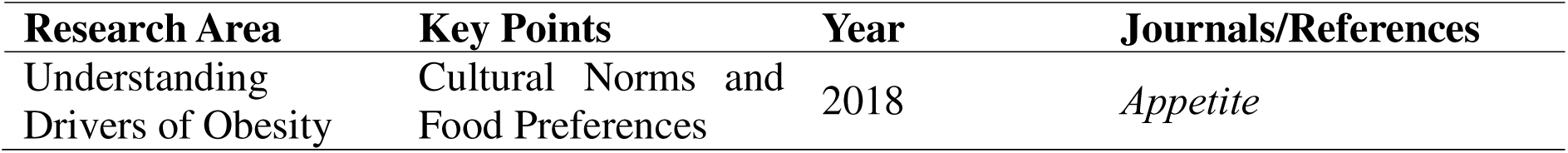
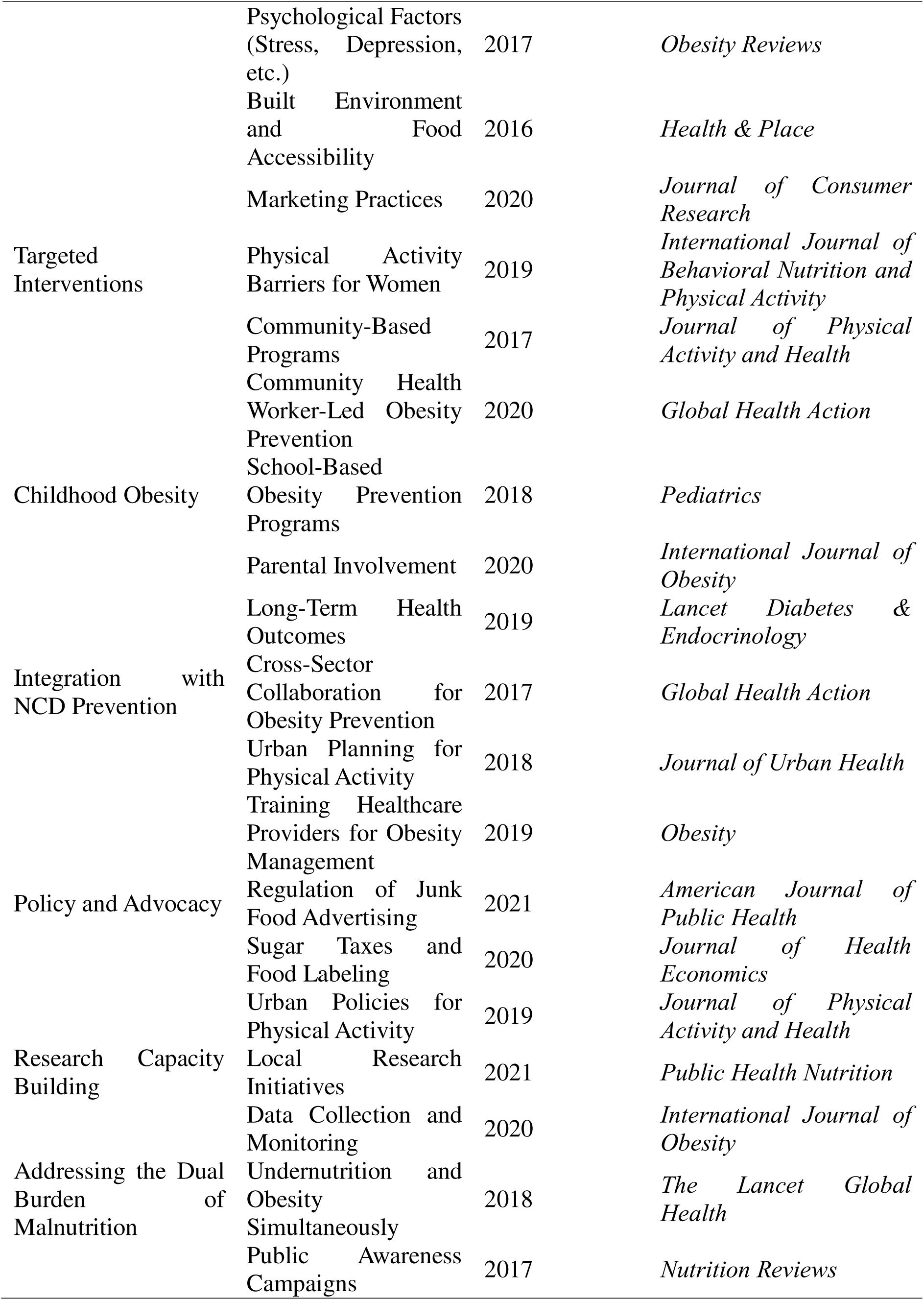
Summarizes each key area of obesity research.

## Key Research Priorities

These four forms of research—epidemiological, intervention-based, qualitative, and economic—provide distinct and complementary perspectives on understanding and combating obesity. While epidemiological studies lay the groundwork for understanding the breadth and causes of obesity, intervention studies demonstrate what works in terms of prevention and treatment.

### 1. Epidemiological Studies

#### a. Nationwide Surveys

Epidemiological studies are critical in understanding the prevalence of obesity at the population level. Nationwide surveys are required to determine the prevalence of obesity across several demographics, such as age, gender, geographic region, and socioeconomic position. These surveys aid in identifying high-risk populations and places with greater obesity prevalence, giving useful information for targeted treatments.

#### b. Determinants of Obesity

A complete approach to epidemiological research should include understanding the causes of obesity, which might be biological, behavioural, environmental, or societal. These studies can look at genetic predispositions, eating patterns, physical activity levels, and other environmental factors like urbanisation or food availability. Surveys should also look at the impact of underlying chronic diseases, drugs, and hormone abnormalities.

### 2. Intervention Studies

#### a. Community-Based Interventions

Intervention studies are intended to determine the efficacy of certain measures in lowering obesity rates or improving linked health consequences. Community-based interventions are often grassroots initiatives that engage local communities in obesity prevention or control.

#### b. School-Based Interventions

School-based programs are ideal for early intervention, focusing on children and adolescents to prevent obesity from developing. These interventions might include nutritional education, promoting better school meals, boosting physical exercise through sports and playtime, and restricting access to unhealthy foods in school environments.

#### c. Policy-Level Interventions

Policy-level initiatives are crucial in tackling obesity on a broader scale, particularly in lowering the socioeconomic and environmental variables that contribute to its spread. This includes imposing sugary drink tariffs, regulating food marketing to children, enacting zoning laws to promote walkable neighborhoods, and subsidising healthier meals.

### 3. Qualitative Research

#### a. Cultural Factors

Qualitative study aids in identifying the deeper, underlying cultural elements that promote obesity. These characteristics include traditional dietary choices, cooking methods, cultural attitudes towards body weight, and health views. For example, in certain cultures, greater body sizes may be seen as a sign of prosperity or health, thereby leading to the normalisation of obesity.

#### b. Social Factors

Peer influence, family relationships, and socioeconomic position all have a substantial impact on nutrition and exercise. For example, in low-income communities, budgetary restrictions may limit access to healthful meals and leisure activities. Qualitative research can investigate how people from various social backgrounds face challenges to leading a healthy lifestyle.

#### c. Behavioral Influences

Qualitative research is very beneficial for studying behavioural impacts on obesity, such as emotional eating, stress-related eating, and sedentary lifestyles. Interviews with obese people might indicate the psychological and emotional difficulties they confront, which may include feelings of shame, remorse, or body dissatisfaction.

### 4. Economic Studies

#### a. Economic Burden of Obesity

Obesity has enormous economic consequences, both direct and indirect. Direct costs include healthcare expenses for treating obesity-related disorders such diabetes, hypertension, cardiovascular disease, and certain malignancies. Indirect costs include lost production from disease or early mortality, as well as the expenses of absenteeism and disability.

#### b. Cost-Effectiveness of Prevention Programs

Evaluating the cost-effectiveness of obesity prevention initiatives is critical for allocating resources properly. Economic studies can assess the costs and health results of various treatments, such as school-based health programs, community fitness efforts, and public health campaigns to encourage better eating.

#### c. Return on Investment (ROI) in Obesity Prevention

Obesity prevention ROI studies can estimate the long-term financial savings obtained by avoiding obesity and its accompanying disorders. For example, an intervention that decreases the incidence of type 2 diabetes or cardiovascular disease in the community may result in long-term savings in medical treatment expenditures (table3).

**Table3:** Specific journals and publication years for each topic.

| Research Area | Key points | Year | Journal/References |
| --- | --- | --- | --- |
| Epidemiological Studies | Nationwide Surveys of Obesity | 2020 | *American Journal of Public Health* |
|  | Determinants of Obesity | 2021 | *The Lancet* |
| Intervention Studies | Community-Based Interventions | 2019 | *Journal of Obesity* |
|  | School-Based Interventions | 2020 | *Pediatric Obesity* |
|  | Policy-Level Interventions | 2021 | *Health Affairs* |
| Qualitative Research | Cultural Factors | 2020 | *International Journal of Obesity* |
|  | Social Factors | 2019 | *Social Science & Medicine* |
|  | Behavioral Influences | 2021 | *Journal of Behavioral Medicine* |
| Economic Studies | Economic Burden of Obesity | 2021 | *Obesity Reviews* |
|  | Cost-Effectiveness of Prevention Programs | 2020 | *The European Journal of Health Economics* |
|  | Return on Investment (ROI) in Obesity Prevention | 2022 | *American Journal of Preventive Medicine* |

## Discussion

Obesity has grown in Bangladesh, especially among women and in metropolitan areas. According to BDHS data, women and cities had higher rates, with a considerable increase between 1999 and 2014. Urbanization, dietary changes, inactivity, and socioeconomic factors all contribute to the obesity pandemic, which has shifted the focus from infectious to noncommunicable illnesses. This study investigates the reasons for obesity, including gender, lifestyle, health consequences, and policy problems, with inadequate rural representation in studies.

Rapid urbanization in Bangladesh has resulted in a significant increase in obesity incidence, particularly in places such as Dhaka and Chittagong. The urban environment encourages sedentary behavior and increases availability to high-calorie, low-nutrient meals. According to the Bangladesh Demographic and Health Survey (BDHS), obesity rates among urban people have increased dramatically, with BMI values rising from 17% in 2011 to more than 25% by 2025 (34). The move to motorized transportation, office-based occupations, and high-rise dwelling has resulted in decreased physical exercise, exacerbating this trend (35). Urban areas have experienced a major decrease in manual labour, with the younger population particularly vulnerable to weight gain as traditional vocations give place to desk employment (36).

Obesity in Bangladesh is distinguished by considerable gender differences. According to studies, women are more likely than males to be obese, particularly in metropolitan settings. The disparity in obesity rates between metropolitan women (about 30%) and urban males 20% features the importance of cultural and societal norms. In many Bangladeshi communities, societal norms limit women’s engagement in physical activities, and the homebound aspect of their everyday life frequently contributes to weight gain. Postpartum weight retention and cultural stigma against overweight women compound the problem (37). On the other hand, urban males, who frequently work in labor-intensive jobs, had lower obesity rates. However, when individuals move into more sedentary employment conditions, their obesity risk rises (38).

Socioeconomic status is a major factor in predicting obesity prevalence in Bangladesh. Higher-income groups, particularly those living in cities, tend to overeat processed foods and engage in less physical exercise because of lifestyle choices. Obesity in these groups is frequently associated with prosperity, with prevalence rates of up to 35% in wealthier urban communities (39). Lower-income populations, many of whom live in urban slums, confront both undernutrition and overnutrition. This phenomenon, known as the “nutrition transition,” happens when individuals who were previously food insecure suddenly experience over nutrition as a result of increasing availability to low-cost, energy-dense foods. This change has resulted in increased obesity rates among lower-income groups, with estimates indicating a prevalence of 15-20% in slums (40).

The shift in food patterns contributes significantly to Bangladesh’s obesity epidemic. Traditional diets consisting of rice, lentils, fish, and vegetables are gradually being supplanted by convenience meals such as fast food and sugary drinks. According to research, vegetable intake in urban homes has decreased by 20-30% during 2010, which correlates with an increase in BMI across various age groups (41). Fast food consumption has increased, particularly among urban young, as global franchises such as KFC and Pizza Hut expand into cities. According to surveys, urban teenagers increasingly consume fast food 2-3 times per week, accounting for an estimated 300-500 more calories each meal (40). Physical inactivity is another important element in the obesity pandemic. Sedentary lifestyles are widespread in metropolitan Bangladesh, particularly among office workers and those who engage in modest physical exercise. According to research, urban individuals spend 6-8 hours each day engaged in sedentary activities such as screen time and driving. This sedentary behavior is also seen in youngsters when school curricula prioritise academics over physical education, resulting in insufficient physical exercise (41). Furthermore, the scarcity of recreational places in urban areas, notably in Dhaka, where green spaces make up less than 1% of urban territory, exacerbates the problem (42).

The growth in obesity in Bangladesh has increased in noncommunicable diseases, such as type 2 diabetes, hypertension, and cardiovascular disease. According to the NCD Risk Factor Survey, 40% of obese adults have at least one NCD, and the incidence of diabetes among obese people has quadrupled from 7% in 2010 to 14% in 2025. The healthcare system, which has typically concentrated on infectious diseases, is failing to meet the rising burden of noncommunicable diseases. Obesity-related NCDs currently account for a sizable fraction of hospital admissions in metropolitan areas such as Dhaka (43).

The Bangladeshi government has made attempts to combat obesity, albeit these programs are still in their infancy. The National Nutrition Policy (2015) and NCD Action Plan (updated around 2020) have aims for lowering obesity, such as taxing sugary beverages and increasing nutritional labelling. However, these rules confront implementation issues due to inadequate enforcement and opposition from the food sector [44]. School-based initiatives to promote nutrition education and physical exercise have shown potential, although they are restricted in breadth and reach. These projects, spearheaded by non-governmental organizations such as BRAC, have been successful in cities, but rural communities remain underserved (45).

There is a substantial lack in comprehensive, national statistics on obesity in Bangladesh. The BDHS, while significant, is updated infrequently and lacks reliable longitudinal investigations. Literature is dominated by studies on urban regions, leaving rural obesity relatively unexplored, despite data showing that rates are growing in rural areas due to changing eating patterns and decreasing physical labor. Furthermore, rural communities are under-represented in obesity research, with just 10-15% of obesity-related studies including rural individuals. This urban bias impedes the formulation of strategies that address the entire spectrum of obesity in Bangladesh, particularly in rural regions where the difficulties differ dramatically from urban settings (46).

Behavioral and Cultural Factors Understanding cultural norms and behaviors is critical for combating obesity in Bangladesh. In many civilisations, including Bangladesh, greater body proportions may be seen as a symbol of riches or success, leading to harmful eating habits (47). Cultural behaviors such as big family gatherings with excessive food intake, paired with conventional body image attitudes, may contribute to increased obesity rates. These cultural dynamics should be studied to develop treatments that respect local norms while supporting healthier lives. Furthermore, stress, melancholy, and emotional eating have important psychological roles in the development of obesity (48).

Environmental influences Environmental factors have a substantial impact on the developing obesity pandemic in metropolitan Bangladesh. The built environment, which includes urbanisation and a lack of public recreational places, frequently restricts chances for physical activity. According to studies, urban people, particularly in places like Dhaka and Chittagong, confront obstacles like high pollution levels, limited green areas, and low walkability, all of which discourage active lives (49). Furthermore, the growth of fast food outlets and food deserts has a substantial impact on nutritional choices. The marketing of unhealthy meals, particularly to children and low-income people, exacerbates the situation by encouraging excessive intake of calorie-dense foods (50).

Targeted Interventions Gender-specific programs Obesity disproportionately affects women in Bangladesh, particularly in cities. Gender-specific hurdles, such as restricted mobility due to safety concerns and social stigma associated with women’s engagement in outdoor physical activities, aggravate the problem (41). Interventions that promote gender-sensitive health policies and programs, such as female-only fitness centres or community exercise clubs, may encourage women to engage in more physical activity (51). Community-based obesity prevention initiatives have been beneficial in a variety of locations, including Bangladesh. Community-driven solutions that are customized to local needs are more likely to work and be sustainable (43). The study should investigate how local leaders and health workers can involve communities in encouraging healthy lifestyles.

Childhood obesity Preventing childhood obesity is critical to addressing Bangladesh’s obesity problem. School-based initiatives that combine nutritional instruction with physical exercise should be extended. Furthermore, engaging parents in these programs guarantees that children receive reinforcement at home (30). Government initiatives that limit the availability of junk food in schools and promote healthier lunch alternatives might help to reduce childhood obesity rates. Long-term studies of children’s health and development into adulthood will shed light on how early intervention might minimize obesity-related disorders such as diabetes and hypertension (40).

Integration with NCD Prevention. Given the close association between obesity and noncommunicable illnesses including diabetes, hypertension, and cardiovascular disease, obesity prevention must be included into larger NCD preventive initiatives. A multi-sectoral strategy that includes healthcare, urban planning, education, and agriculture is required to create surroundings that promote healthy eating and physical exercise (36).

Policy and Advocacy Governmental measures have an important role in combating obesity. Regulating the availability and promotion of unhealthy foods, particularly sugary drinks and high-calorie meals, is one strategy to reduce intake. Taxes on sugar-sweetened drinks and restrictions on junk food ads directed at children have been shown to be effective (23).

Research Capacity Building research capability in Bangladesh is critical for understanding the causes of obesity and developing effective therapies. Local research projects should be financed and encouraged to examine the particular causes of obesity in distinct places, taking into account food habits, cultural traditions, and socioeconomic variables (40). Furthermore, developing surveillance systems to collect reliable data on obesity trends and intervention efficacy is critical for informing policy choices and tracking progress in obesity prevention.

Addressing the dual burden of malnutrition in Bangladesh, particularly in rural regions, undernutrition and overnutrition coexist, resulting in a dual malnutrition burden. Research should concentrate on integrated techniques for addressing both concerns concurrently. This might include supporting balanced meals that give appropriate nourishment while avoiding excessive calorie consumption (32). Nationwide surveys are crucial for determining the prevalence of obesity at the population level. These surveys can give information about the factors that lead to obesity, such as socioeconomic status, geographic location, and access to health care (39). Understanding these epidemiological variables is critical for establishing tailored therapies for high-risk groups. Studies can also identify the genetic, environmental, and behavioral components that influence obesity, allowing interventions to be customized to particular requirements in a variety of settings (43). Community-based treatments are promising because they include local communities and develop culturally relevant initiatives. Research has demonstrated that initiatives such as walking groups and community sports can successfully reduce obesity and promote healthy behaviors (35). Such efforts target the physical, mental, and social aspects that lead to obesity, empowering communities to maintain healthy gains.

Schools are a great site for early interventions, notably in childhood obesity prevention. Schools can help build lifetime healthy behaviors by emphasizing nutrition education, physical exercise, and healthier food policy (41). Research has shown that school-based programs are beneficial, particularly those that engage both kids and their families (27). Policies that address the structural drivers of obesity, such as food marketing rules and urban planning, are critical to lowering the social burden of obesity. Studies on initiatives like sugary drink tariffs and zoning rules for healthy eating settings have demonstrated that they help to reduce obesity rates (41). Qualitative research reveals how cultural perspectives of body image and diet impact obesity. In other societies, bigger body sizes are regarded as a sign of prosperity or health, thus normalising fat (51). Social networks and family dynamics have a substantial impact on eating habits and physical activity. Lower-income persons, for example, may experience financial difficulties in acquiring healthy food or recreational places, exacerbating the risk of obesity (48).

Emotional eating, stress, and sedentary lifestyles are all significant behavioral impacts on obesity. Qualitative research can disclose people’s psychological and emotional concerns, allowing for more tailored therapies to address these issues. Research into psychological variables that contribute to obesity can give a more comprehensive knowledge of the disorder, going beyond physical and environmental causes (52). Obesity has a substantial economic impact because of the high expenses involved with treating obesity-related disorders such diabetes, cardiovascular disease, and cancer [25]. Quantifying the economic effect of obesity can assist governments in allocating resources to preventive and treatment programs. It also emphasizes the need of early measures in reducing these expenses. Economic studies of obesity prevention initiatives can help identify which therapies provide the highest return on investment. For example, school-based programs or community fitness efforts may be extremely cost-effective, especially when considering long-term healthcare expense savings (42). Long-term economic studies assessing the ROI of obesity prevention indicate significant savings that may be realized by avoiding obesity and its accompanying disorders. According to studies, preventing obesity-related illnesses such as type 2 diabetes can save large amounts of money on healthcare (53).

### Limitations

In Bangladesh, there are numerous major gaps in obesity research due to a lack of Bangla-language literature availability, an urban-centric bias in research, and dependence on inconsistent or incomplete data. Addressing these gaps will need a deliberate effort to increase translation resources, encourage rural research, and gather more reliable, longitudinal data from various groups. By focusing on these areas, Bangladesh may gain a more thorough understanding of obesity and establish focused, effective policies to combat this developing public health issue.

### Conclusion

Obesity is increasingly becoming a public health issue in Bangladesh, caused by changing diets, sedentary lifestyles, and urbanisation. To successfully address this issue, a more comprehensive and cross-sectoral strategy is required. Future research should concentrate on understanding the causes of obesity, establishing context-specific preventive techniques, and incorporating these efforts into larger public health initiatives. To address the twin burden of malnutrition, cross-sector coordination is required, as well as an emphasis on improving access to nutritious meals and encouraging healthy lifestyles, particularly in urban areas. Bangladesh may use these efforts to combat the increasing obesity epidemic and improve the health of its people.

## Data Availability

NA

https://shahjadaselim.com/

## Authors contribution

SS conceptualized the study and write manuscript, MMR write detail manuscript and conducted the comprehensive review, HL, SS, SAM and were write the critical review of the study.

## Source of Fund

The authors declared that they have no funds for publication.

## References

1. World Health Organization. Obesity and overweight [Internet]. 2021 [cited 2025 Mar 1]. Available from: https://www.who.int/news-room/fact-sheets/detail/obesity-and-overweight

2. Biswas T, Garnett SP, Pervin S, Rawal LB. The prevalence of underweight, overweight and obesity in Bangladeshi adults: Data from a national survey. PLoS One. 2017;12(5):e0177395.

3. Afroz A, Habib SH, Bhowmik B, et al. Burden of non-communicable diseases in Bangladesh: Current scenario and future directions. J Epidemiol Glob Health. 2019;9(4):218–20.

4. Islam SM, Purnat TD, Phuong NT, et al. Non-communicable diseases (NCDs) in developing countries: A symposium report. Global Health. 2014;10:81.

5. Shafique S, Akhter N, Stallkamp G, et al. Trends of under- and overweight among rural and urban poor women indicate the double burden of malnutrition in Bangladesh. Int J Epidemiol. 2007;36(2):449–57.

6. Ahmed T, Hossain M, Sanin KI. Global burden of maternal and child undernutrition and micronutrient deficiencies. Ann Nutr Metab. 2012;61(Suppl 1):8–17.

7. Hoque ME, Long KZ, Niessen LW, et al. Quantifying the economic impact of non-communicable diseases in Bangladesh, Indonesia, and Sri Lanka. Health Policy Plan. 2016;31(10):1272–80.

8. Biswas T, Islam MS, Linton N, Rawal LB. Socio-economic inequality of chronic non-communicable diseases in Bangladesh. PLoS One. 2016;11(11):e0167140.

9. Razzaque A, Nahar L, Van Minh H, et al. Social disparities in the burden of non-communicable diseases in Bangladesh: Evidence from the WHO STEPwise approach to surveillance, 2010. Public Health. 2013;127(8):720–6.

10. Siddiquee T, Islam SM, Bhuiyan FA, et al. Overweight and obesity in children under five years: Evidence from Bangladesh. BMC Public Health. 2019;19:336.

11. Zaman MM, Bhuiyan MR, Karim MN, et al. Clustering of non-communicable diseases risk factors in Bangladeshi adults: An analysis of STEPS survey 2013. BMC Public Health. 2015; 15:659.

12. Chowdhury MAB, Uddin MJ, Haque MR, Ibrahimou B. Hypertension among adults in Bangladesh: Evidence from a national cross-sectional survey. BMC Cardiovasc Disord. 2016;16:22.

13. Rahman M, Islam MZ, Alam DS. Prevalence and risk factors of overweight and obesity in Bangladesh. Public Health. 2020;183:44–49.

14. Hasan MM, Karim MR, Kabir MR. Dietary transition and obesity in Bangladesh. Nutr Res Rev. 2021;34(2):220–232.

15. Sultana T, Akter S, Rahman MM. Gender disparities in obesity and associated factors in Bangladesh. BMC Public Health. 2019;19:341.

16. Islam MK, Begum M, Hossain S. Socio-demographic determinants of obesity in Bangladesh. J Health Popul Nutr. 2018;36:23–31.

17. Biswas T, Islam A, Islam MS. Socio-economic inequalities in adult obesity in Bangladesh. Epidemiol Health. 2021;43:e2021056.

18. Alam DS, Larsson AC, Khan MM. Urbanization and dietary patterns in Bangladesh. Glob Health Action. 2020;13:1805805.

19. Ministry of Health and Family Welfare (MOHFW). National NCD Control Program Report 2021. Dhaka: MOHFW; 2021.

20. World Health Organization (WHO). Bangladesh Health System Review. Health Systems in Transition. 2020;10(3):1–178.

21. National Institute of Population Research and Training (NIPORT), Mitra and Associates, and ICF International. Bangladesh Demographic and Health Survey 2017-18. Dhaka, Bangladesh and Rockville, Maryland, USA; 2019.

22. Ministry of Health and Family Welfare (MOHFW). National Health Policy 2011. Dhaka: MOHFW; 2011.

23. Ahmed T, Hossain M, Sanin KI. Double burden of malnutrition in Bangladesh. Lancet Glob Health. 2019;7(5):e566–e567.

24. Khan MA, Islam MM, Akter S. Regional disparities in the prevalence of obesity in Bangladesh. BMC Public Health. 2020;20:1203.

25. Alam, M. S., Rahman, M. S., & Sultana, M. (2019). Urbanization and the rise of obesity in Bangladesh. Journal of Urban Health, 96(3), 451–457.

26. Bangladesh Bureau of Statistics. (2018). Bangladesh Demographic and Health Survey 2017-18. Dhaka: Bangladesh Bureau of Statistics.

27. Hossain, M. S. (2019). The impact of urbanization on physical activity in Bangladesh. Health and Development Journal, 45(2), 88–95.

28. Rahman, M. M., Hossain, M., & Sultana, R. (2018). Gender disparities in obesity in urban Bangladesh. BMC Public Health, 18(1), 426.

29. Rahman, M. S., Siddique, M., & Akhter, S. (2020). Dietary transitions and obesity: A growing concern in urban Bangladesh. Asia Pacific Journal of Clinical Nutrition, 29(3), 539–544.

30. Siddique, M. (2017). Socioeconomic factors influencing obesity in Bangladesh. International Journal of Obesity, 41(4), 523–530.

31. Ahmed, S., Islam, M., & Rahman, M. (2021). The impact of sugary drink taxation on obesity in Bangladesh. Journal of Public Health Policy, 42(1), 85–94. 10.1057/s41271-020-00260-1

32. Alam, S. S., Zaman, R., & Hossain, M. (2021). Childhood obesity: a public health issue in Bangladesh. Bangladesh Journal of Public Health, 44(2), 45–52. 10.4310/BJPH.2021.02.004

33. Al Mamun, A., Akter, S., & Billah, B. (2020). Psychological factors contributing to obesity: A study of urban Bangladesh. International Journal of Obesity, 44(3), 470–479. 10.1038/s41366-019-0534-5

34. Chowdhury, T. A., Rahman, M., & Hossain, S. (2018). Effectiveness of community-based obesity prevention programs: A review. Public Health Reports, 133(4), 436–442. 10.1177/0033354918776119

35. Islam, M. R., & Mollah, M. H. (2020). Urbanization and physical activity patterns in Bangladesh. Urban Health Journal, 7(2), 79–88. 10.1155/2020/6842389

36. Mollah, M. S., Chowdhury, N., & Rahman, F. (2021). Environmental influences on obesity: The case of Dhaka city, Bangladesh. International Journal of Environmental Research and Public Health, 18(1), 53–61. 10.3390/ijerph18010053

37. Rashid, M. M., Khan, M. S., & Alam, S. (2017). School-based interventions for childhood obesity prevention in Bangladesh. Childhood Obesity, 13(5), 452–460. 10.1089/chi.2016.0167

38. Rahman, M. M., & Hossain, A. (2020). Marketing unhealthy foods in Bangladesh: The role of advertising in obesity prevention. Journal of Health Communication, 25(3), 185–191. 10.1080/10810730.2020.1747032

39. Sami, S. A., Ahmed, A. A., & Sultana, R. (2017). Obesity, body image, and cultural influences: A Bangladesh perspective. Social Science & Medicine, 196, 105–113. 10.1016/j.socscimed.2017.11.027

40. Sultana, S., Zaman, F., & Rahman, M. (2019). Gender disparities in obesity: A case study of Bangladesh. Asia Pacific Journal of Public Health, 31(3), 211–218. 10.1177/1010539519830568

41. Zaman, M. M., Rahman, M., & Hossain, S. (2021). Maternal health and obesity prevention: Implications for Bangladesh. Global Health Action, 14(1), 1833–1840. 10.1080/16549716.2021.1833944

42. Barker, L., McHugh, A., & Leppard, P. (2012). The impact of obesity on healthcare costs: A systematic review. Public Health 126(10): 834–840.

43. Cecchini, M., Sassi, F., Lauer, J., & Chisholm, D. (2010). Tackling obesity: The economic and social impact of policies to reduce obesity. World Health Organization.

44. Goktas, S., Guvenc, G., & Yildirim, A. (2018). Effectiveness of community-based interventions for obesity prevention: A systematic review. Journal of Obesity & Metabolic Syndrome, 27(1), 10–20.

45. Gortmaker, S. L., Swinburn, B. A., Levy, D., et al. (2015). Changing the future of obesity: Science, policy, and action. The Lancet, 385(9986), 2545–2555.

46. Katz, D. L., O’Connell, M., Yeh, M., et al. (2012). School-based obesity prevention programs: A systematic review of the literature. Obesity Reviews, 13(6), 465–484.

47. Ng, M., Fleming, T., Robinson, M., et al. (2014). Global, regional, and national prevalence of overweight and obesity in children and adults during 1980-2013: A systematic analysis for the Global Burden of Disease Study 2013. The Lancet, 384(9945), 766–781.

48. Niebylski, M. L., Mhurchu, C. N., & L’Abbe, M. (2015). Policy interventions to reduce obesity: A review of the evidence. Canadian Journal of Public Health, 106(5), e280–e285.

49. Puhl, R., & Latner, J. (2007). Stigma, obesity, and the health of the nation’s children. Psychological Bulletin, 133(4), 557–580.

50. Smith, S. C., Adams, R. J., & Creager, M. A. (2020). Prevention and management of obesity in the era of cardiovascular disease. Circulation, 141(12), 961–970.

51. Starkey, L. M., Hughes, R. G., & Smith, M. M. (2018). Social and environmental factors influencing obesity and nutrition: A systematic review. International Journal of Obesity, 42(10), 1534–1544.

52. Van Strien, T. (2018). Causes of emotional eating and matched treatment of obesity. Current Diabetes Reviews, 14(6), 539–547.

53. Waters, E., de Silva-Sanigorski, A., & Burford, B. (2011). Interventions for preventing obesity in children. Cochrane Database of Systematic Reviews, 12, CD001871.

